# Assessment of Donated Medications and their Use in Tikur Anbessa Specialized Hospital: Promoting Green Health and Circular Economy

**DOI:** 10.1101/2024.04.24.24306299

**Authors:** Elbethel Getinet Alemaw, Bezawit Negash Demisse

## Abstract

**Introduction:** Despite the increased access to medicine, developing countries receive large amounts of medicines from donations. Donated medication could save lives and alleviate suffering but they could also cause deleterious effects on recipients. These effects can lead to the disposal of harmful drugs, affecting health infrastructure and causing negative. Organizations donate drugs for various reasons, including charitable, publicity, or humanitarian reasons. Despite the need for donated drugs, drug dumping remains a prevalent issue in Sub-Saharan Africa.

The volume of pharmaceutical waste that accumulates worldwide is now a major concern. The idea of a sustainable, circular bioeconomy on a global scale is appealing. It prioritizes environmental sustainability and human well-being atop economic recovery.

The primary objective of this study was to evaluate the status of donated medicines in Tikur Anbessa Specialized Hospital (TASH) and develop suggestions. Medicines donated from April 2022 up to May 2022 were included in this study

**Materials and Methods:** A cross-sectional survey conducted at an institutional level employed both quantitative and qualitative research methodologies. The study was conducted from April 2022 to May 2022, focusing on pharmaceuticals donated to Tikur Anbessa Specialized Hospital (TASH) between November 2021 and March 2022. For the qualitative part, employees of Tikur Anbessa Specialized Hospital (TASH) working at different dispensary units, and pharmaceutical stores constituted the source population.

**Results and recommendations:** A total of six donors contributed a combined value of 11,715,389 Ethiopian birr worth of medicines, supplies, and equipment. The donated items comprised 45 product categories and a total of 160 individual products. Among these, the most prevalent product categories included Opioids (27.48%), ASRVI (10.22%), PDE-5 Inhibitors (7.70%), Antihemophilic agents (7.34%), Antibiotics (7.02%), Anticonvulsants (5.67%), and Antifungals (3.36%). Management of medicines, supplies, and equipment at Tikur Anbessa Specialized Hospital (TASH) is facilitated through the Pharmaceutical Management Information System (PMIS), similar to other medication management processes. The acquisition of donated medications primarily occurs through both recipient requests and donor initiatives. Delivery of drugs near expiry, too small amount, discontinuation, disposal cost and push pull scenario were among the problems. Simple effective communication among donors and the hospital can uplift these problems and enhance the positive impacts of donations. Furthermore, the hospital needs to update its policy on donation and schedule donations to optimize the effects of donations; as a result promoting sustainability.

## • Introduction

The world’s population has access to medicine more than ever before, albeit with substantial disparities. More than half of the world population consumes more than one dose per day of medicine. In 2020 only 4.5 trillion doses have been given, accounting for $1.4 trillion. This is a 24% increase from 2015. ^1^ However, third-world countries only produce 7% of the drugs they consume.^2^ Thus, they rely on donated drugs for the provision of essential medicines for the majority of their population. ^3^ This demand overshoots in the face of disasters and suffering.

Nevertheless, medication wastage is a significant public health problem in third-world countries. Donated drugs could be inappropriate or unsuitable for treating diseases of the recipient. Most medical donations; arrive near expiration dates and are sent without asking recipients about their needs or prior notifications.^4^ In order to alleviate this problem and optimize the positive impact of donated drugs, WHO developed 12 guidelines in 1999. ^5^ However, the positive impact of donated drugs is affected by proper storage, transportation, and dispensing. ^6^ The level of handling and management of donated drugs are, thus, detrimental.^7^

Despite concerted efforts to build universal formularies to manage pharmaceuticals in developing countries, shipment of brand-name products, expired products, and "garbage bags" containing a disparity of sample drugs in two-tablet packaging still prevails.^8^ Moreover, natural disasters and wars in developing countries creates opportunities to dump unwanted, expired, dangerous products into the affected areas. ^9^

WHO has been advocating optimum communication between donors and recipient countries, as donors should not send drugs the receiver doesn’t need.^5^ However, this effort is hindered by the lack of good governance. ^10^ The political nature of access to medication as well as intense lobbying by all principal stakeholders, makes guidelines impractical in third-world setup.^11^

Donated drugs can positively impact and alleviate suffering or stifle health and worsen it. ^12,13^ Mostly in Ethiopia, extent of the donations is not quantified, and analyzed thus not prioritized. Evidences on the proper execution of these donated medicines, their appropriateness, expiry date, and whether they are demand-driven or not, are also lacking. This study is among the first studies to analyze donated drug conditions at a tertiary level public hospital in Ethiopia. It will highlight donated drug practices and gaps of donating practices in Ethiopia. Hence, data generated from this study will be use to provide recommendations for institutional practices. It will shed light on key points in optimizing donation practices. The findings of this study can direct government and other stakeholders to strategize concerted effort in optimizing drug donation practices.

Essential medicines can only save lives and improve health when they are available, affordable, of assured quality, and properly used. ^14^ Hence, donated drugs during a crisis or as a long-term aid can contribute in provision of affordable essential medicine. For instance, billions of tablets, capsules, and intravenous and oral solutions have been donated, along with the manufacturing, supply chains, and research necessary in an effort to overcome the burden of Neglected Tropical Diseases (NTD). This resulted in significant decline in the prevalence of NTD.^13^

On the contrary, drug donation can be used as a way of dumping certain groups’ drugs, which have deleterious effects. The cost of disposing such medical waste will further debilitate the health infrastructure. Such cases were seen in Sri Lanka, Sudan, and Bosnia. ^11,15,16^ Organizations donate drugs for a variety of purposes. Some may seek tax deductions for charitable purposes; some may seek positive publicity; some may want to dispose of unwanted products without having to pay for destruction; others may do it for humanitarian reasons.^8^ Despite the substantial need of donated drugs in Sub Saharan region drug dumping is a dominant pattern. ^16^Several expired, banned/toxic products, useless, ambiguous, and non-informative, were dumped in large quantities as an international response to the call for help by the Sudanese government and Mozambique. ^12,16^

The volume of pharmaceutical waste that accumulates worldwide is now a major concern. Pharmaceutical waste can come from a number of sources, such as unused, spilled, expired, or contaminated pharmaceuticals^17^. Since medicines are necessary for maintaining human health, they play a significant role in achieving the Sustainable Development Goal (SDG) of "Good Health and Well-Being" set forth by the UN. In the meantime, 4.4% of global greenhouse gas emissions are attributable to the healthcare sector, which includes medicines^18^. Improved management of medications can minimize waste, cut expenses, and lessen adverse environmental effects by adhering to the principles of the circular economy. Adding a cyclical donation process to pharmaceuticals can improve sustainability. Applying circular concepts to reduce pharmaceutical waste is a critical role for pharmacists and healthcare organizations^17^. The idea of a sustainable, circular bioeconomy on a global scale is appealing. It prioritizes environmental sustainability and human well-being atop economic recovery^19^. Although the current guidelines are a good starting point, data suggests that they should be updated to reflect the problems with drug contributions today^20^.

The Objectives of the Study was to assess the status of donated medicines in TASH specifically, to characterize the management of donated medications in Tikur Anbessa Specialized Hospital and to exploring the practice and perception of managers involved in the management of donated medications.

## • Methodology

### • Study Setting

Tikur Anbessa specialized hospital is the largest referral hospital in the country with 200 doctors, 379 nurses, and 115 other health professionals. Undergraduate medical students and residents in the School of Medicine also provide patient care. It has 950 permanent and contract administration staff to support the activities.^6^

#### 2.2. Study Design and Period

The study period was from April 2022 to May 2022 An Institutional based cross-sectional survey that employs quantitative and qualitative research approaches was used.

#### 2.3. Sampling and Sample Size Determination

The study period was from April 2022 to May 2022. All pharmaceuticals that have been donated from November 2021 up to March 2022 at Tikur Anbessa Specialized Hospital (TASH) were analyzed. For the qualitative part, employees of TASH working at different dispensary units as managers and dispensaries, and store keepers were the source population. All complete TASH registry records were used as source data. Due to inconsistencies in the recording system, recent 4 months of receiving vouchers were used.

For the key informant interview purposive sampling was used. Saturation point was reached at the tenth interview, however for assurance purposes two more key informants were interviewed.

### • Data collection procedure

Both primary and secondary data was used for this research; primary data from interview and secondary data from TASH registry records were used.

The data abstraction tool used in this study was adapted from a similar study conducted in Tanzania. ^2^ Data collection was done completely by the investigator and questionnaires were developed in Amharic version and back translated to English. The investigator recorded and transcribed the interview.

## • Method of Data analysis

Collected data was entered into excel and exported into SPSS (Version 26). For quantitative data descriptive analysis such frequency, percentage, mean, and standard deviation were used. Transcribed data was qualitatively described in light of prior literatures and investigators experience.

## • Results

### • Donors, Costs of Donations, and Shares

During this period, 11,715,389 birr worth of medicines, supplies and equipment were donated. The value of the donation made by the organization6 was not indicated, and hence the cost of donations is likely higher than indicated. Highest donations (64 %) was donated by organization 1 but contributed 38.22% of total value (Figure 1 and 2). The total value of donations was highest for organization 4 which only donated once but contributed 57% to the total value.

**Figure 1:** Frequency and Share of donors to total donation

**Figure 2:** Percentage of frequency of donation of medicines donated by each donor

**Figure 3:** Percentage of donations by total value

**Figure 4:** Problems Associated with donated programs

**Table 1:** Total cost of donations made by donors.

| **Donor Organization** | **Total Cost of Donation in Birr** | **Percentage** |
| --- | --- | --- |
| Org 5 | 113,349 | 0.96 |
| Org 3 | 14,820 | 0.13 |
| Org 2 | 4,477,731 | 38.22 |
| Org 1 | 413,083 | 3.53 |
| Org 4 | 66,96,406 | 57.16 |
| Org 6 | 0 | 0 |
| Total | 11,715,389 | 100 |

### • Product Categories

There were 45 donated product categories and 160 products donated. Opioid, ASRVI, PDE-5 Inhibitor, Antihemophilic agent Antibiotic, Anticonvulsant, and Antifungal accounted for 27.48 %, 10.22%, 7.70 %, 7.34%, 7.02 %, 5.67% and 3.36%. Each product category has different medicines and hence time that was left for expiry was only available for a product. Some product categories such as Acetylcholinesterase, Analgesic, Anti-convulsant, Anti arrhythmic, ARBs and General Anesthesia has only one product. Twenty-one product categories had only one product and 12 have two products. ARVI has the most products (33 products), followed by Antibiotic (17 products) (Table 2).

**Table 2:** Product Category and Quantity of various donated medicines.

| Product Category | No of Product | Quantity | Percentage of product |
| --- | --- | --- | --- |
| Acetylcholinesterase | 1 | 500 | 1.01 |
| Analgesic | 1 | 800 | 1.62 |
| Anti-convulsant | 1 | 200 | 0.41 |
| Anti-arrhythmic | 1 | 1225 | 2.48 |
| Anti-emetic | 1 | 500 | 1.01 |
| ARVI | 4 | 103 | 0.21 |
| Antibiotic | 17 | 3465 | 7.02 |
| Anticholinergic | 1 | 660 | 1.34 |
| Anticonvulsant | 4 | 2800 | 5.67 |
| Antiemetic | 1 | 600 | 1.22 |
| Anti-fungal | 4 | 1660 | 3.36 |
| Antihemophilic agent | 12 | 3620 | 7.34 |
| Anti-Histamine | 1 | 500 | 1.01 |
| Anti-Malarial | 2 | 1330 | 2.70 |
| Antineoplastic | 2 | 210 | 0.43 |
| ARBs | 1 | 500 | 1.01 |
| ARVI | 33 | 5042 | 10.22 |
| General Anesthesia | 1 | 40 | 0.08 |
| Benzodiazepine | 3 | 100 | 0.20 |
| Beta blocker | 2 | 40 | 0.08 |
| Birth Control | 1 | 2 | 0.00 |
| Calcium channel blocker | 1 | 50 | 0.10 |
| Calcium Supp | 1 | 12 | 0.02 |
| Cardiac glycoside | 2 | 70 | 0.14 |
| Combination antibiotic | 2 | 80 | 0.16 |
| Disinfectant | 1 | 40 | 0.08 |
| General anesthetic | 1 | 23 | 0.05 |
| H2 Blocker | 2 | 30 | 0.06 |
| Implantable contraceptive | 1 | 100 | 0.20 |
| ab regent | 2 | 40 | 0.08 |
| Laxative | 1 | 15 | 0.03 |
| Local Anesthesia | 4 | 103 | 0.21 |
| Neuromuscular blocker | 2 | 146 | 0.30 |
| NSAID | 2 | 244 | 0.49 |
| Obstetrics and Gynecological medication | 6 | 367 | 0.74 |
| Opioid | 8 | 13561 | 27.48 |
| Oral hypoglycemic drug | 1 | 4 | 0.08 |
| PDE-5 Inhibitor | 2 | 3800 | 7.70 |
| potassium supplement | 2 | 5110 | 10.36 |
| PPI | 3 | 93 | 0.19 |
| Statins | 1 | 30 | 0.06 |
| Supplement | 1 | 30 | 2.82 |
| Thiazide Diuretics | 2 | 23 | 5.61 |
| Vaccine | 15 | 1065 | 2.16 |
| Vitamins | 3 | 410 | 0.83 |

### • Survey on Donated Medicines, Supplies and Equipment to TASH

Even though the exact number of donated medicines, equipment or supplies cannot be estimated simply,91.7% of responders indicated that it is possible to have a reasonable estimate by counting the receipt. One respondent said “TASH will receive about 1 billion birr estimated donated medicine and supplies pharmaceuticals per year”.

Most respondents (83.3%) indicated that once received, the donated medicine equipment’s or supplies are managed through TASH information system as any other medicines, supplies or equipment. When interviewed about the management system one respondent said “These donated products are not managed by a dedicated supply chain channel team. They are managed together like other pharmaceuticals. For these donated pharmaceuticals Model is cut separately.

TASH uses PMIS (Pharmaceutical management information system) to handle its medications. There is also a manual method of managing the supplied products”.

Donations at TASH follow two systems. The first system is donation programs that secure continuous supply for ART, and anti -TB, anti-malarial from dedicated donors. On the other hand, donations from international/ local donors (both institutional & individual donors) occur for a brief duration. It is facilitated by a dedicated team from the hospital that sends letter of request to the donors. Upon receiving of the donation certificate, the donors arrive with listed items. Local donors, however, can either use normal distribution pathways or supply directly to the hospital. A respondent said, “There are two types of donation systems. The first is programmed donation system in which there are continuous suppliers. The program pharmaceuticals such as ART, oncologic, anti-TB and anti-malaria drugs are continuously available in this hospital. The other type is donation which the donation from other health institutions or from international missions and individual donors. There is established team in the hospital that communicates and sends request to the donors to obtain the pharmaceuticals”. Another interviewee also said, ” There is established team in the hospital that communicates with the donors to obtain the pharmaceuticals. If the source of donated pharmaceuticals is from hospitals the hospital staff sends the request to the donors and then the pharmacist cut model 19 and receive the donated items. The drug then distributed to pharmacy unit and dispensed for free. If the donated item is medical equipment the letter is written for the intended purpose. The donors receive a Donation Certificate and come with item list after they have cleared customs. It comes with an item list and an import license that permits you to negotiate with the custom authorities. Following clearance, these donations are transported to the hospital”. A respondent from oncology department also said, “They donate every month once they start giving. On occasion, cancer specialist doctors take the initiative and speak with donors regarding required medications”.

These drugs are detrimental especially to those with financial limitations, as they can acquire them for free. An interviewee highlighted this by saying, “TASH makes use of a variety of medications and medical devices that are donated from sources. The health-care system is able to function because of the numerous donations available. The donated items will be able to meet the needs of a wide variety of hospital patients. The hospital does not have a plan in place for accepting donations because it is relying on the motivation and initiative of the volunteer donors. Drugs not available in the country like chemotherapy drugs can be lifesaving to many. Moreover, they are very expensive and only received through donations, enabling TASH to provide cancer treatment to patients. One interviewee said,” It is usually leukemia drugs that enter the hospital in the form of donations. We do not send a list of medicines, but the donors themselves send us a list and donate by their intention”. Another one also indicated how lifesaving these donations are by saying,” Drugs that are not available in the national supply chain may be donated to the hospital in certain situations via donation only. These medications, in turn, play an important role in preventing significant drug access problems in patients”.

These donations could be obtained through request by the recipient (8.3%) alongside initiation of the donor (91.7%). The donors either make their donations directly to the hospital on a regular basis or supply based on the need of the hospital. They send a request detailing what pharmaceuticals drugs and supplies the hospital requires and in what quantity. Then the donation ensues. A respondent said, “TASH receives pharmaceutical products from donors who have expressed an interest in doing so. They might be advertising their product indirectly in this scenario. When TASH runs out of medications or supplies, the team will contact donors and request what they need. There is no clear plan for what products and how much of them the hospital will need at any one time to meet the demands of the patients”.

The donation is not a response to a request by the hospital. However, some of such donors do communicate with the hospital on the need. “Most of the donors once start giving their contribution they donate every month. On occasion, cancer specialists take the initiative and communicate with donors regarding medications required by the hospital”. The government might be a proxy receiver to the donation in some cases that distributes to hospitals needing donations.

There is always that chance donation might be wasted. One respondent explained instances how this happens by saying, “Quantification of the received product wasted by expiration requires full information in all hospital pharmacy units so that it is difficult to determine. Some of the items distributed to different units may not be used properly because their expiry date will be too short. They will start expiring at anywhere at store, ward area and dispensary area with in short period of receiving these items”. Those with short expiry dates may not last longer. Even though it is difficult to quantify wasted donations (50%), the wastage is significant in most cases (41.7%). One respondent explained this by saying, “It’s possible that the drugs provided are really not what the hospital needs. If unneeded pharmaceuticals are sent in, they will be expired, and raising the hospital’s and country’s disposal costs. They go to the hospital to aid, but there are times when there aren’t many unneeded donations. Despite the fact that numerous drugs are needed, donors can only provide what they have or can donate. There is also an issue is one of transportation in which the Pharmaceuticals with short half-life frequently expire in a short period of time after reaching the hospital due to the long travel time”. Problems associated with donated medicine are multiple. Donated medicines could be near to expiry dates (33.3%). As one respondent said, “The expiration rate of donated products is higher than other purchased medications and supplies”. They might be too small (16.7%) or get discontinued (8.3%) at times. Donated medicines and supplies can incur disposal cost (8.3%). In rare cases unwanted medicines might be supplied (8.3%). What’s more, companies could enforce to purchase some other item from the institution in order to donate. This push pull mechanism could incur unnecessary cost (25%), one respondent among others who mentioned this said, “Push system is the major challenging problem which is enforcing the hospital to buy additional pharmaceutical together with the donated item. The additional bought pharmaceutical may not be as such important to the hospital. Lack of previous data for appropriate estimation of required pharmaceuticals is another problem”. As another respondent put it, “Some of the suppliers may donate or distribute their near to expire product via push system. This system enforces the receivers (in this case TASH) to have bulk product which will be prone to wastage”.

If communication and distribution channels are optimized these problems could have been averted as timely delivery of drugs is solely dependent on simple and effective communication. A respondent among others who suggested this said, “In order to reduce wastage and shortage of pharmaceutical in the hospital, there could be improved communication between donors and acceptors. Instead of simply pushing their donation, donor organizations should assess the hospitals’ need for their products”. The communication shall also be among institutions as bulk delivery can be relieved by channeling to hospitals in short supply of the donation. One respondent when asked for suggestion as to how problems can be solved replied, “Creating a system that does not rely on the push system. Receiving things based on hospital usage data. It is possible to distribute these products to other health facilities that need it if they arrive in bulk with an end date that is too short”. Accurate estimation of the need based on historical data on the receiver end is vital to obtain donation in appropriate quantity. A respondent suggested this by saying, “Accurate estimation of required drug and supplies based on consumption data is required for obtaining them in appropriate quantity. This prevents wastage or shortage of donation items after reaching the hospital”. Improving the policy (83%) as well as scheduling donations (25%) were believed to resolve the problems by the respondents. A respondent among the others who mentioned this said, “Because there are so many distribution channels between them, making optimal use of donated medicinal supplies is problematic. Too longer process to get these items into the hospital makes may reduce the wise use of donations. If not available with in short period of time these items will expire before consumption”. Besides the avoidance of push pull system could help significantly (8.3%). A respondent explained how this system highly affects wastage as follows,” If possible the hospital staff could record how much donation received each time from licensed donors. The donors shouldn’t attach the crucial items with unimportant items via push system of donation. It is preferable not to offer drugs that the donors are aware will expire soon”. Some respondents underlined that donation should not be overlooked, but there is a need for further study on this aspect. A respondent suggested more research and quantification can be a solution by saying,” Accurate estimation of required drug and supplies based on consumption data is required for obtaining them in appropriate quantity”. At the end of an interview a respondent commented by saying,” Regarding to pharmaceutical donations there should be further study that study about:

It should be the responsibility of the receiver (the hospital) to determine how many medications are required. This is due to the fact that donors may not be fully aware of the needs of recipients. The relationship between donors and recipients of donations should be thoroughly studied and established”.

The major strength of the study lies in its incorporation of both qualitative and quantitative study designs. It has tried to give a descriptive analysis of the prevailing donation practices at TASH and its underlying problem. It is however limited by design to identify determinant of such donation practices. As these is a descriptive study actual association between factors cannot be sought.

## • Discussion

In developing countries like Ethiopia where donors cover 41% of the health expenditure contribution of donated medicine equipment and supplies is detrimental to survival. ^7,21^ This has been shown in the ongoing pandemic of Covid 19. They have aided in saving lives and controlling the pandemic which would have otherwise been catastrophic. Similarly, the continuous donation of anti-cancer medication to TASH has enabled TASH to provide curative and rehabilitative treatment to cancer patients across the country. These would have not been possible without donation as such medications are very expensive even in western setup. The donations to TASH were polarized as they are dominated by two companies. Donations were based on demand and its donation management practices are similar to the nationwide practice of donated medicines. The donations are dominated by two donors that supply either on a continuous basis or once from international and/ or local donors. Mostly such donations are initiated by both the donor and the receiver.

Unlike the case of Sudan & Bosnia, drug dumping was not witnessed in our setup as the expiry date of drugs donated ranged from 46 – 62 months year. ^12,14,16,22^ But still donation of near expiry date drugs and too small donations compared to the demand were seen alongside unanticipated discontinuation of donation. Push pull mechanisms by suppliers were also seen. This forces the receiver to purchase another item for the donation to be secured resulting in unnecessary cost.

The necessity of efficient communication is implied in order to optimize donation and minimize problems; as a result promoting sustainability. Accurate estimate of the need by the receiver based on experience eases the communication and enables delivery of appropriate amount and type of donation. Consequently decreasing the adverse effect pharmaceutical waste can have on our environment. This is similar to the experience seen in Uganda where communication has fostered a positive outcome from donations.

## Conclusions

Donated medicines, supplies, and equipment has immensely contributed to the management of patients in TASH. They have alleviated suffering for many lives, which wouldn’t have been otherwise impossible. Donations at times has been hijacked by institutions as a method of advertisement or marketing strategy (push pull). But mostly drugs donated are far from expiry date and based on necessity decreasing unnecessary disposal costs. Henceforth, experience of TASH on donation can be used as a benchmark for effective donation practices.

Problems of drug donations and negative impacts on the environment due to wastage can also be minimized by improving the policy of in the receiver end and scheduling donations as to optimize their timely delivery to patients. Further research is needed to assess the determinant of donation practices from donor and receiver end so as to give a more founded recommendation that can achieve a more sustainable and greener healthcare system and utilize the concept of circular economy.

## Declarations

### Ethical approval and consent to participate

Ethical approval was obtained from the ethical review committee of Addis Ababa University, School of Pharmacy with reference number (ERB/442/14/2022). Written informed consent was obtained from all study participants before the questionnaire was administered and collected data were kept confidential.

### Consent for publication

‘Not applicable’

### Availability of data and materials

The datasets generated and analyzed during the current study are not publicly available due to privacy and ethical concerns but are available from the corresponding author upon reasonable request.

### Competing interests

The authors declare that they have no competing interests.

### Funding

‘Not applicable’

### Authors’ contributions

B.N. and E.G. drafted the proposal work, synthesized the appropriate methodology, conducted the data analysis, data collection, generated the results, and write-up of the study. All authors read and approved the final manuscript.

## Data Availability

No legal or Ethical restrictions.

## Acknowledgments

The authors would like to thank Addis Ababa University College of Health Science and the staff of Tikur Anbessa Specialized for their unlimited and kind help throughout the data collection process.

## Notes

### Competing Interest Statement

The authors have declared no competing interest.

### Clinical Trial

Not Applicable

### Funding Statement

We did not receive any external funding or financial support during the course of this research. The study was conducted independently, and all data collection, analysis, and manuscript preparation were carried out without any financial assistance.

### Author Declarations

Ethical approval and consent to participate Ethical approval was obtained from the ethical review committee of Addis Ababa University, School of Pharmacy with reference number (ERB/442/14/2022). Written informed consent was obtained from all study participants before the questionnaire was administered and collected data were kept confidential.

